# Optimal Timing of Organs-at-Risk-Sparing Adaptive Radiation Therapy for Head-and-Neck Cancer under Re-planning Resource Constraints

**DOI:** 10.1101/2024.04.01.24305163

**Authors:** Fatemeh Nosrat, Cem Dede, Lucas B. McCullum, Raul Garcia, Abdallah S. R. Mohamed, Jacob G. Scott, James E. Bates, Brigid A. McDonald, Kareem A. Wahid, Mohamed A. Naser, Renjie He, Aysenur Karagoz, Amy C. Moreno, Lisanne V. van Dijk, Kristy K. Brock, Jolien Heukelom, Seyedmohammadhossein Hosseinian, Mehdi Hemmati, Andrew J. Schaefer, Clifton D. Fuller

## Abstract

**Background and Purpose:** Prior work on adaptive organ-at-risk (OAR)-sparing radiation therapy has typically reported outcomes based on fixed-number or fixed-interval re-planning, which represent one-size-fits-all approaches and do not account for the variable progression of individual patients’ toxicities. The purpose of this study was to determine the personalized optimal timing for re-planning in adaptive OAR-sparing radiation therapy, considering limited re-planning resources, for patients with head and neck cancer (HNC).

**Materials and Methods:** A novel Markov decision process (MDP) model was developed to determine optimal timing of re-planning based on the patient’s expected toxicity, characterized by normal tissue complication probability (NTCP), for four toxicities. The MDP parameters were derived from a dataset comprising 52 HNC patients treated at the University of Texas MD Anderson Cancer Center between 2007 and 2013. Kernel density estimation was used to smooth the sample distributions. Optimal re-planning strategies were obtained when the permissible number of re-plans throughout the treatment was limited to 1, 2, and 3, respectively.

**Results:** The MDP (optimal) solution recommended re-planning when the difference between planned and actual NTCPs (ΔNTCP) was greater than or equal to 1%, 2%, 2%, and 4% at treatment fractions 10, 15, 20, and 25, respectively, exhibiting a temporally increasing pattern. The ΔNTCP thresholds remained constant across the number of re-planning allowances (1, 2, and 3).

**Conclusion:** In limited-resource settings that impeded high-frequency adaptations, ΔNTCP thresholds obtained from an MDP model could derive optimal timing of re-planning to minimize the likelihood of treatment toxicities.

**Graphical Abstract:** Overview of the analysis method. The sub-figures displaying Organs at risk and toxicities are adapted from [1] with permission. Abbreviation: NTCP = Normal tissue complications probability.

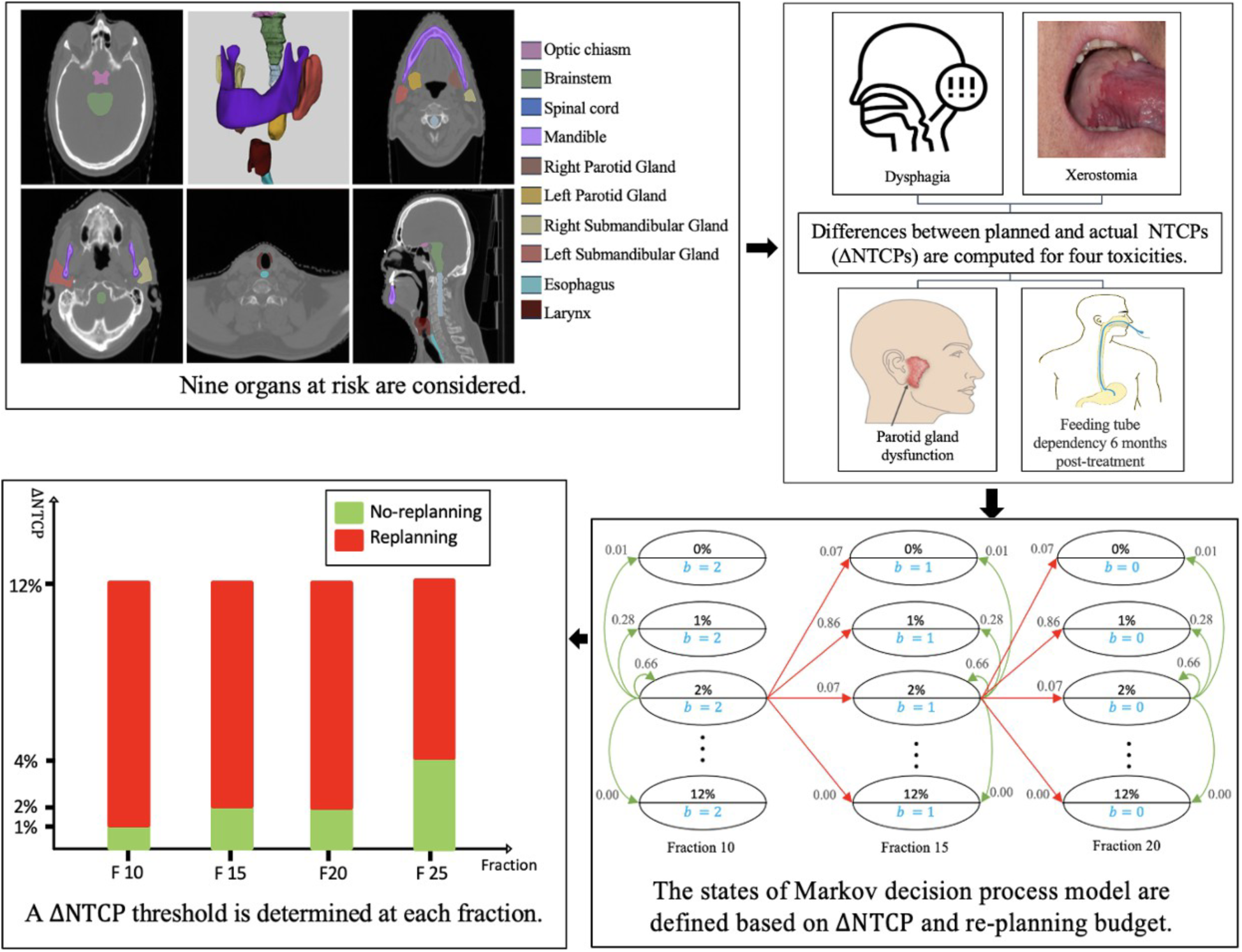

## 1 Introduction

Advancements in radiation delivery techniques, such as intensity-modulated radiation therapy (IMRT) and volumetric-modulated arc therapy, enable accurate dose delivery to tumor targets while minimizing radiation exposure of the surrounding organs at risk (OARs) [1]. However, anatomical changes during the treatment, such as weight loss or tumor shrinkage, may cause the actual delivered dose to OARs to deviate from the planned dose. This can increase the risk of treatment-induced toxicities, particularly in cases where multiple OARs are in close proximity to the target, as in head and neck cancer (HNC) [2–4]. To address this, adaptive radiation therapy (ART) has been clinically introduced, proposing on-therapy re-planning in response to anatomical changes in the target and OARs [5–10].

In practice, however, the clinical implementation of ART with daily (or even less-frequent) re- planning remains limited, in large part due to the extensive human/personnel/workflow resources required to frequently perform key tasks such as segmentation and quality assurance as well as limited device accessibility time [11, 12]. Recent artificial intelligence (AI)-based algorithms (such as auto-segmentation or synthetically created CTs) [13, 14] may mitigate some or all of these process level frictions; however, the integration of such AI tools within the ART workflow is still evolving [15]. With the advent of hybrid MR-Linac devices, real-time adjustment of daily radiation plans, known as on-line ART, is now a possibility. On-line ART can also be enabled with the availability of high-frequency, high-quality cone- beam CT or CT-on-Rails devices [16, 17]. Regardless of ART implementation imaging inputs (MR or CT), cancer centers in the U.S. typically have implemented ART at fixed intervals, notably once mid-therapy [18] and often as a ‘verification’ of re-simulation. Most of the relevant studies also only report outcomes on fixed-number and/or fixed-interval re- planning [19–21] (see Supplementary Table A1 for a comprehensive literature review). Such pre- determined schedules for treatment re-planning, however, take a one-size-fits-all approach and do not account for the uncertain trajectory of individual patients’ toxicities [21], nor patient-specific tumor regression. As a result, determining the optimal timing of re-planning episodes remains a crucial unmet need, particularly for OAR-sparing adaptive approaches (whether for MR-Linac as we have implemented in MR-guided clinical trials [19] or for analogous CT-based approaches [20]).

Heukelom et al. [22] investigated the optimal implementation of ART with a single re-planning allowance (in OAR-sparing radiation therapy) using daily on-treatment CT imaging with a CT-on-rails device. Leveraging the same dataset, this paper presents a new analytical approach to derive optimal re-planning strategies based on Markov decision process (MDP) models. Our aim is to identify the optimal timing for re-planning based on changes in normal tissue complication probabilities (NTCP) of four toxicities: xerostomia, dysphagia, parotid gland dysfunction, and feeding tube dependency at 6 months post-treatment. We further include allowances in HNC treatment plan adaptations (through limiting the number of available re-plans) to enhance personalized treatment and efficacy. MDPs constitute a class of mathematical optimization models that aim to determine optimal decisions/actions in stochastic dynamic systems [23, 24]. MDPs have been successfully employed to find the optimal timing for various medical interventions [25–31]; however, to our knowledge, MDPs have not been applied for triggering adaptive re-planning. We develop a generalized framework for the utilization of MDPs for evidence-based individualized radiation treatment re-planning, scalable across resource-rich and resource-limited facilities, and applicable to both CT- and MR-based platforms. Thus, rather than a class solution based on population estimates of toxicity reduction potential, we enable personalized adaptive therapy with consideration of a budget.

## 2 Materials and Methods

### 2.1 Data

This study used a prior dataset of CT-on-Rails image-guided radiation therapy (IGRT), detailed by Heukelom et al. [22], which comprised information from patients treated for HNC at the University of Texas MD Anderson Cancer Center between 2007 and 2013; this retrospective secondary analysis was performed under MD Anderson Cancer Center Institutional Review Board approval (MDA RCR03-0800). The treatments involved (chemo-) radiotherapy with daily CT-on-Rails IGRT. Of the 52 patients, 36 were male and 16 were female. Among them, 46 patients were aged 18-65, while the remaining 6 were older than 65. The primary cancer sites were as follows: Larynx (1 patient), Oropharynx (13), Oral cavity (4), Hypopharynx (0), Nasopharynx (15), and Sinonasal (12). Treatment modalities included radiotherapy alone (16 patients), induction chemotherapy followed by radiotherapy (2), induction chemotherapy followed by concurrent chemoradiation (16), concurrent chemoradiation (14), and radiation plus Cetuximab (4). The patients’ characteristics are summarized in Supplementary Table B1.

For these 52 HNC patients, Heukelom et al. [22] calculated deviation of the actual dose from the planned dose for nine OARs at fractions 10 and 15 of the treatment. At each fraction, they estimated NTCP for the toxicities related to the OARs (xerostomia, dysphagia, parotid gland dysfunction, and tube feeding dependency at 6 months post-treatment) by projecting the actual dose through the remainder of the treatment period. Subsequently, they compared these findings with the planned NTCPs and determined the difference, i.e., ΔNTCP, for each toxicity. The NTCP models are presented in Supplementary Table E1. The MDP model presented in this paper used the ΔNTCP from this dataset [22], which are summarized in Table 1. For each observed ΔNTCP value, Heukelom et al. [22] reported the number of patients for whom this ΔNTCP was the highest value among the four NTCP models.

**Table 1.**
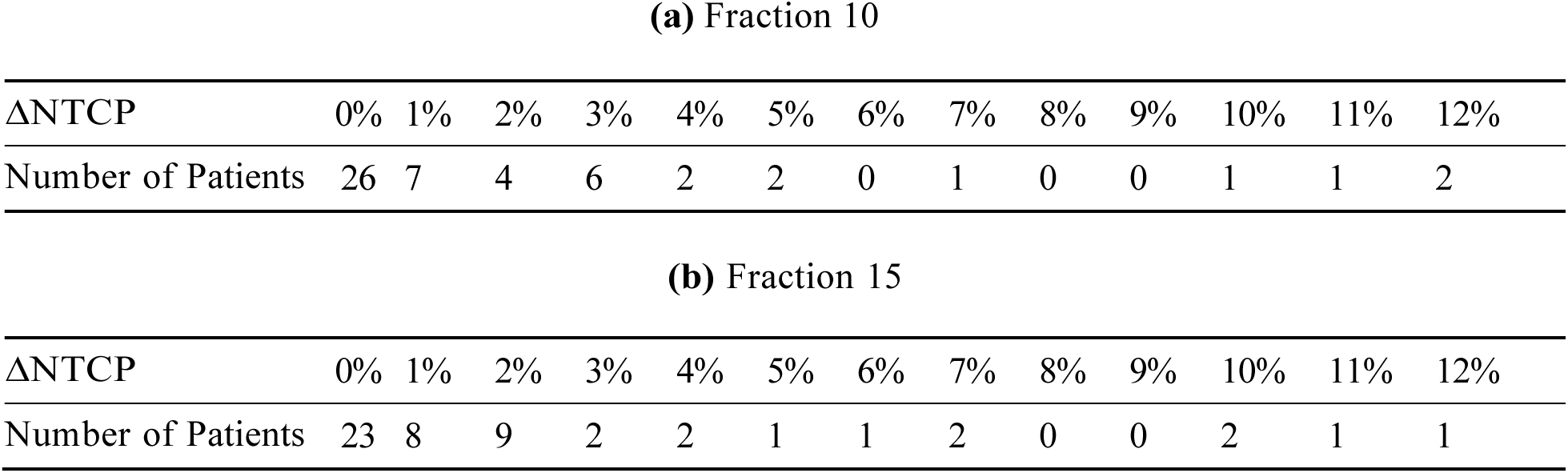
Observed ΔNTCP values based on the difference between planned dose and actual dose, along with the number of patients associated with each ΔNTCP value. Adapted from [22] with permission.

*2.2 Decision Model*

In the MDP model, estimates of an individual patient’s toxicity outcome, as a function of the delivered radiation dose to OARs, determine the state of the system at each decision epoch during the treatment (e.g., day). Depending on the observed state, the clinician may decide between two possible actions: (1) Re-plan or (2) continue with the current plan. When the action is to continue with the current plan, the system may transition from one toxicity state to another stochastically, governed by transition probabilities. Re-planning changes the probabilistic transition towards more favorable outcomes/states. Given that a limited number of ‘re-planning’ actions may be taken throughout the treatment, an optimal solution to the MDP identifies the optimal timing for taking such actions, as a function of the toxicity states. The MDP model captures the stochastic evolution of post-treatment toxicity risk and identifies optimal re-planning times to mitigate the toxicities (if necessary). The model components are as follows:

#### Re-planning allowance

Depending on available resources for plan adaptations, the model considered a maximum number of re-plans *B* that could be implemented throughout the treatment. The analysis was performed for *B* = 1, 2, 3.

#### Decision epochs

Given a treatment period consisting of 33-35 fractions, the decision epochs were set at fractions 10, 15, 20, and 25. Prior studies have shown that anatomical changes are unlikely to happen very early during the treatment [22]; thus fraction 5 was omitted. Fraction 30 was also excluded due to its proximity to the end of treatment, with negligible impact on the total dose to the OARs.

#### States

At each decision epoch, the state of the system was captured by the pair (ΔNTCP*, b*), where ΔNTCP denoted the deviation of treatment toxicity from the planned value at that time, and *b ≤ B* was the number of remaining re-plans. For example, if the maximum permissible number of re-plans was 2 (*B* = 2), and the clinician opted to implement a re-plan at fraction 10, then the state of the system at fraction 15 became *b* = 1. The ΔNTCP ranged from 0% to 12% in the model (Table 1); in computing the number of cases for each reported ΔNTCP value, only those patients were included who experienced the change of ΔNTCP in at least one of the four aforementioned toxicities.

#### Actions

At each decision epoch, two possible actions were included: (1) Re-planning, or (2) continuing with the current plan (no re-planning). The stochastic transition of the toxicity state from one decision epoch to the next was a function of the action taken and was determined by the associated transition probabilities. Fig. 1 illustrates the states of the MDP and possible transitions associated with each action at Fractions 10, 15, and 20 along with their smoothed transition probabilities, for the case that at most 2 re-plannings may be performed.

**Fig. 1:**
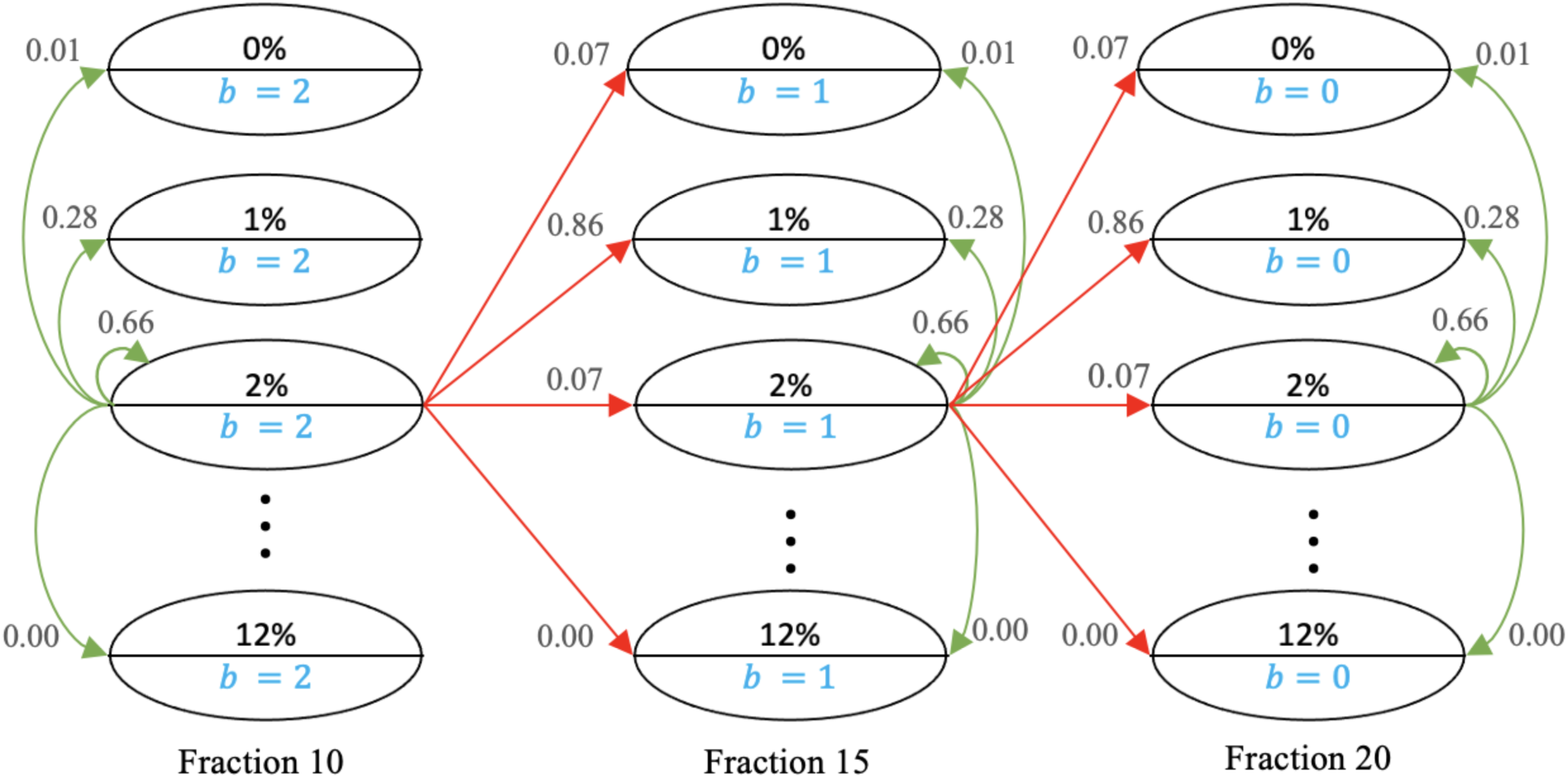
Markov Decision Process Model. The permissible number of re-plans is 2 (*B* = 2). The states are shown by ellipses.,6.NTCP values are located in the upper half of the ellipses, while the lower halves contain the value of *b* (the number of remaining re-plans). Transitions between states are shown by green arrows when the action is ‘no re-planning’ and by red arrows when the action is ‘re-planning.’ Not all arrows, states, and fractions are included to avoid ambiguity. For example, the smoothed transition probability from state (2%,2) state to (1%,1) when the action is ‘re-planning’ is 0.86.

#### Transition Probabilities

The transition probabilities, governing the stochastic evolution of toxicity under each action, were estimated using the information presented in Table 1. Under the ‘no re- planning’ action, the probabilities for transitions from fraction 0 to 10 and from fraction 10 to 15 were directly estimated based on the number of patients in each ΔNTCP category. For example, the probability of transitioning from ΔNTCP = 0% at fraction 0 to ΔNTCP = 1% at fraction 10 is 7*/*52 = 0.13, as 7 patients out of 52 exhibited such a change in ΔNTCP from fraction 0 to fraction 10. To make our transition probabilities more realistic with a broader spread and to avoid any deterministic transitions between states, we smoothed out the transition probability matrix from Fraction 10 to 15 by using kernel density estimation with Gaussian kernels, implemented in Python [27]. We manually varied the bandwidth between 0 and 1 to observe its effect on the distribution. Based on these tests, we selected a final bandwidth of 0.4, balancing smoothness and fit to capture meaningful data patterns without introducing noise. The original and smoothed probabilities associated with the ‘no re-planning’ action are presented in Supplementary Material C. It was assumed that transition probabilities from fraction 10 to 15 remained constant for subsequent decision epochs, due to the absence of reported ΔNTCP information beyond fraction 15 by Heukelom et al. [22]. To model the impact of the ‘re-planning’ action on transition probabilities, it was assumed that, at each decision epoch, the action immediately decreased ΔNTCP to a value proportional to the elapsed treatment time. For example, if the ‘re-planning’ action was taken at fraction 10 for ΔNTCP = 5%, the state of toxicity would immediately decreased to ΔNTCP = 2% at this fraction, as approximately one third of the treatment period had passed and the dose escalation would not continue through the rest of the treatment; see Fig. 2. Subsequently, the transition from fraction 10 to 15 was governed by the probabilities for ΔNTCP = 2%, calculated from Table 1. The probabilities associated with the ‘re-planning’ action, as described above, are presented in Supplementary Material C.

**Fig. 2:**
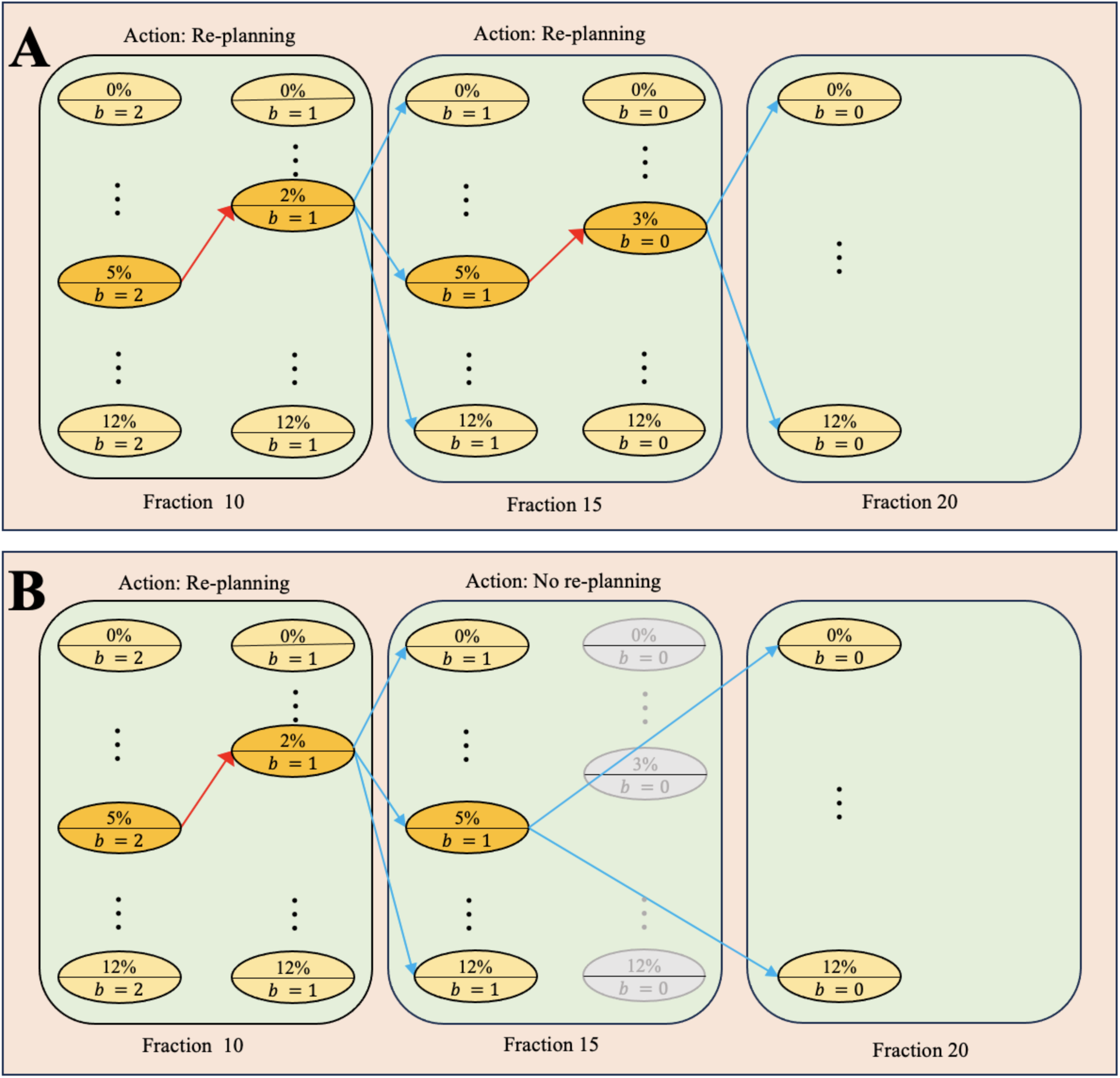
Effect of actions on system transitions. The permissible number of re-plans is 2 (*B* = 2). The states are represented by ellipses. The,6.NTCP values are located in the upper halves of the ellipses, while the lower halves contain the value of *b* (the number of remaining re-plans). When the action at a fraction is ‘re-planning,’ as in Fractions 10 and 15 in Part A and Fraction 10 in Part B, the transition to a state with a lower,6.NTCP within the same fraction is shown with red arrows. Transitions between states from one fraction to the next are shown with blue arrows. When the action is ‘no re-planning,’ as in Fraction 15 in Part B, no transition occurs within that fraction. Instead, the system transitions to a new state at Fraction 20. This is depicted by the gray states at Fraction 15, indicating that there is no immediate decrease in,6.NTCP.

#### Rewards

For each set of consecutive actions taken at the decision epoch, the model considered the expected −ΔNTCP at the end of the treatment period (with respect to the transition probabilities) as the corresponding reward. The objective of the MDP was to maximize the expected reward by identifying an optimal set of actions, one at each decision epoch, as a function of the system’s state. This is referred to as an optimal policy. Because the rewards were defined by negative values in the MDP model, smaller end-treatment ΔNTCP values translated to higher rewards.

The optimal policy of an MDP may become a single-threshold policy (also referred to as control-limit policies) [24], which refers to a class of policies that use thresholds on the state value to recommend an action. For this study, a single-threshold policy recommended re-planning when the ΔNTCP exceeded a threshold, while no re-planning was needed when it falls below the threshold. This implies that if the action for ΔNTCP = *x*% at a certain decision epoch was to re- plan, then for every other state with ΔNTCP = *y*% *> x*% and the same allowance *b*, the action was also to re-plan. A single-threshold policy reduces the complexity of decision-making to a simple rule that uses only one threshold at each fraction to trigger action and is efficient to implement.

The MDP was solved using the MDPtoolbox of MATLAB [32], for *B* = 1, 2, 3. The MATLAB code and its outputs are available at https://figshare.com/s/ 64bc3481737d17fc287e.

## 3 Results

After solving the MDP, the optimal policy, which specified the optimal action (‘re-planning’ or ‘no re-planning’) for each state (ΔNTCP, *b*) at each decision epoch, was reported. The analysis revealed that the optimal policy was a single-threshold policy, where a specific ΔNTCP threshold was assigned to each fraction. When only one re-plan was allowed (*B* = 1), the optimal policy at fraction 10 was to re-plan for any ΔNTCP value greater than or equal to 1%. Subsequently, at fraction 15, this threshold increased to 2% and remained at 2% for fraction 20. At fraction 25, the minimum ΔNTCP required for a re-planning was 4%. These thresholds remained the same in the optimal policies for *B* = 2, 3. We summarized the results in Table 2 and provided an illustration in Supplementary Fig. D1 for *B* = 3.

**Table 2.** Optimal re-planning thresholds based on ΔNTCP and re-planning allowance.

| Re-planning allowance ($B$) | 1 | 2 | | 3 | | |
| --- | --- | --- | --- | --- | --- | --- |
| Number of remaining re-plans ($b$) | 1 | 2 | 1 | 3 | 2 | 1 |
| ΔNTCP threshold at fraction 10 | 1% | 1% | - | 1% | - | - |
| ΔNTCP threshold at fraction 15 | 2% | 2% | 2% | 2% | 2% | - |
| ΔNTCP threshold at fraction 20 | 2% | 2% | 2% | 2% | 2% | 2% |
| ΔNTCP threshold at fraction 25 | 4% | 4% | 4% | 4% | 4% | 4% |

## 4 Discussion

This study introduced the first — to our knowledge — application of the mathematically rigorous MDP methodology, to determine optimal timing of ART in HNC. The MDP model guides clinicians in determining the minimum values of ΔNTCP at fractions 10, 15, 20, and 25 for performing a re-plan, given a re-planning allowance of 1, 2, or 3 throughout the treatment.

The re-planning ΔNTCP threshold increased over time in the optimal policy, consistent with the diminishing impact of re-planning as the treatment progresses. These values were 1% at fraction 10, 2% at fractions 15 and 20, and 4% at fraction 25. Additionally, the results suggest optimality of re-planning for any changes in NTCP (ΔNTCP *≥* 1) at fraction 10. This supports the findings of Heukelom et al. [22], who identified fraction 10 as the optimal time for a single re-plan. Importantly, at a given fraction, the ΔNTCP thresholds remained the same for different number of re-plans (*B* = 1, 2, 3); the re-planning allowance did not affect these thresholds. Furthermore, in cases where ΔNTCP at fraction 25 was below the 4% threshold, both options (‘re-planning’ or ‘no re-planning’) yielded the same impact on the end-treatment ΔNTCP, indicating that re-planning did not result in an improvement.

We acknowledge that minimizing the expected ΔNTCP may result in prescribing re-planning for any ΔNTCP value, potentially leading to a high number of false negatives. This arises from our modeling assumptions, which permitted a fixed number of re-plans at no cost, encouraging frequent re-planning due to low ΔNTCP thresholds. In an ongoing study, we are incorporating re- planning costs without limiting the number of re-plans to better explore the trade-off between cost and benefit; however, at a minimum, we have opted to err on the side of patient benefit, rather than cost- control; as patient NTCP benefit is potentially scalable across any health system, while cost per re-plan and acceptable cost constraints are variable across national and international health policy and reimbursement systems.

The significance of a ΔNTCP of 4% or below lies in its potential impact on patient outcomes. While seemingly minor, even slight decreases in NTCP can have considerable implications for patient health and well-being. For instance, a reduction in severe toxicity such as osteoradionecrosis by just 4% could significantly enhance the quality of life and long-term survival prospects for patients. Moreover, customizing treatment to achieve such reductions underscores the importance of personalized care tailored to individual patient needs. By focusing on optimizing treatment outcomes at this level, healthcare practitioners can prioritize patient-centric approaches that aim to minimize treatment-related toxicity and maximize overall patient benefit.

We leveraged an existing CT-on-rails reference dataset [22] to objectively derive the ΔNTCP listed in the proposed MDP model. It is crucial to acknowledge the limitations regarding the generalizability of findings from this single-site retrospective in silico dataset. For instance, the in silico daily dose accumulation was not actively applied to individual patients, but rather calculated post hoc from a high-granularity CT-on-rails daily volumetric IGRT series. The CT-on-rails platform at MD Anderson utilized an in-house custom- constructed intermediary localization and alienation software (CT-Assisted Targeting (CAT)) [33]. Consequently, there were instances where delivered geometric shifts were either unrecorded or unrecoverable, or clearly aberrant (such as extensive shift records representing an initial setup that was then revised after repositioning) during the secondary export of coordinate displacement to the commercial Record and Verify software (Mosaiq, Elekta AB). These discrepancies were subsequently omitted in the in-silico model to streamline data, leading to conceptual gaps in the resultant NTCP modeling where these missing values were not accounted for. Furthermore, it is important to note that this modeled secondary dataset did not include adaptation or daily re-optimization of the initial daily dose in vivo. Consequently, the data presented in this paper should be viewed as a clinically approximate semi-synthetic illustrative use-case, rather than a definitive rationale for the large-scale implementation of the observed idealized re-planning thresholds across distinct operational platforms. Nonetheless, we believe that the resultant MDP model could be readily scaled using higher-quality prospective or observational cohort data for secondary validation. In essence, the data presented in this paper should be regarded as informative rather than definitive, and the individualized planning parameters suggested by MDP should be viewed as proof of concept rather than a formal criterion barring external validation.

In the prior work, Heukelom et al. [22] exclusively reported ΔNTCP for fractions 10 and 15, consistent with internal re-planning practices at MD Anderson Cancer Center based on data from a Phase II study by Maki et al. [34]. Consequently, for model extensibility in the current application, we have explicitly assumed transition probabilities remain stable for the subsequent epochs. This assumption introduced a known level of uncertainty that warrants consideration and is an area of future research, as it has been unclear for specific OARs whether these transition states were indeed stable over therapy. Moreover, the MDP model stipulated weekly re-planning intervals on indexed fractions (e.g., fractions 10, 15, 20, 25, and 30) as a simplification for clarity of presentation reflective of our current adaptive protocols [19], but could readily be adapted to continuous daily fraction-based re-planning intervals.

The four NTCP models in Supplementary Table E1 are among the most currently used models to calculate the NTCP values for the considered toxicities: xerostomia, dysphagia, parotid gland dysfunction, and feeding tube dependency at 6 months post-treatment [22, 35]. With the advent of more recent NTCP models for HNC radiation therapy, e.g., [36], it is possible that new NTCP models could offer improved estimations. In addition, the presented results are based on data collected from 52 HNC patients. While we recognize the importance of sample size in calibrating MDPs, this sample size is considered substantial in HNC research, considering that HNC accounts for only about 4% of cancer cases in the United States [37]. Furthermore, our results are contingent upon the available CT-on-Rails data, and future research may benefit from incorporating higher-dimensional data (e.g., GTV/the clinical target volume (CTV) modifying approaches, MRI anatomic and/or biomarker data for TCP/NTCP) for a more extensive insight.

Nonetheless, the proposed MDP model for ART is clinically relevant, mathematically rigorous, resource-aware, and scalable, and can be adjusted based on new OAR toxicity with reference NTCP values. Necessarily, the precision of the model relies on accurate calculations of NTCP, particularly when adhering to rigorous criteria that determine whether patients are suitable for or excluded from ART [38]. Despite the challenges and limitations, our study introduces novel contributions to the field of ART. Unlike previous works [22], which primarily considered a single re-planning allowance, our optimization model extends its applicability to scenarios with multiple re-plans. Furthermore, our optimal policy spans across fractions 10, 15, 20, and 25, representing an advancement beyond the limited scope of Heukelom et al. [22], which only reported results for fractions 10 and 15. Gan et al. [39] achieved optimal timing for re-planning in HNC radiation therapy by analyzing weekly dose data through semi-auto segmentation and the K-nearest-neighbor method. They constructed a dose deviation map to visualize differences between planned and actual doses, simulating different ART scenarios. By evaluating accumulated dose differences before and after re-planning, the optimal timing for re-planning was determined. Our methodology differs in that we incorporate ΔNTCP and utilize it to develop an MDP model for determining the optimal timing for re-planning.

An aspect not explored in this study is the adaptation based on GTV or CTV modification, either for shrinking GTV/CTVs [19] or isotoxic boost approaches [40, 41]; we concentrated solely on OAR-based adaptation. Adapting based on GTV could open avenues for optimal re-plans, potentially influencing NTCP and extending into scenarios such as Stereotactic Body RT [42]. This introduces a distinctive problem and solution space beyond the scope of our current investigation. Furthermore, we exclusively focused on optimizing the ART workflow within the context of photon therapy. Similar optimization methodologies could prove advantageous when exploring ART in the context of proton therapy, particularly in addressing setup variability reduction [43]. For this study, NTCP calculations were based on the ‘plan of the day.’ While our findings may vary with deformable dose registration, the operational implementation remains consistent. Future efforts should consider incorporating deformable dose registration to enhance the model’s generalization.

Several surveys in both low/middle-income countries [44, 45] and high-income economies [45] have identified resource constraints as an impediment to ART implementation; the use of models such as our MDP provides a potential avenue for stratification of resource allocation. Put simply, with one re-plan allowed, almost all patients would be best served via re-planning early during treatment; however, as the budget of re-planning staff/technical/time resources expand, evidence-based personalized re-planning is potentiated by our MDP approach.

## Data availability statement

The anonymized data from the enclosed manuscript has been deposited at DOI:10.6084/m9.figshare.25517338; a referee version is available for review at https://figshare.com/s/64bc3481737d17fc287e.

## Preprint availability

A preprint/pre-peer review version of the enclosed manuscript has been submitted in accordance with NIH NOT-OD-17-050, “Reporting Preprints and Other Interim Research Products” simultaneous with initial submission for peer-review and is available at https://doi.org/10.1101/2024.04.01.24305163.

## Acknowledgment

This work was supported by the National Institutes of Health (NIH) National Cancer Institute (NCI) Research Grant (R01CA257814), the NCI Cancer Center Support Grant Program in Image-Driven Biologically-informed Therapy (IDBT) Program (P30CA016672), and the Image Guided Cancer Therapy Research Program at The University of Texas MD Anderson Cancer Center, and the MD Anderson Charles and Daneen Stiefel Center for Head and Neck Cancer Oropharyngeal Cancer Research Program. Lucas B. McCullum and Raul Garcia were supported by the NCI Supplement program under R01CA257814-02S2, and R01CA257814-03S2, respectively. Drs. McDonald and Wahid are both supported by the Image-guided Cancer Therapy T32 Fellowship (T32CA261856). Dr. Heukelom received related support from the Netherlands Rene Vogels Fond Fellowship. Drs. Heukelom and van Dijk received relevant support from the Dutch Cancer Society/KWF Kankerbestrijding. Dr. Fuller received related support from the National Institute of Dental and Craniofacial Research (NIDCR) (R01DE028290). Dr. Fuller has received related direct industry grant/in-kind support, honoraria, and travel funding from Elekta AB. Dr. Fuller has served in an unrelated consulting capacity for Varian/Siemens Healthineers. Philips Medical Systems, and Oncospace, Inc. Dr. Brock received unrelated support from the Helen Black Image Guided Fund, RaySearch Laboratories AB, the Apache Corporation, and the Tumor Measurement Initiative through the MD Anderson Strategic Initiative Development Program (STRIDE) and various NIH mechanisms. Dr. Scott received relevant support under the NCI Case Comprehensive Cancer Center Support Grant (P30CA043703), with additional unrelated NIH support. Dr. Moreno received related support from NIDCR (K01DE030524) and NCI (K12CA088084) during the project period, with additional unrelated NIH support.

## Supplementary Material A. Literature Review on Head-and-neck-cancer Adaptive Radiation Therapy

**Table A1:**
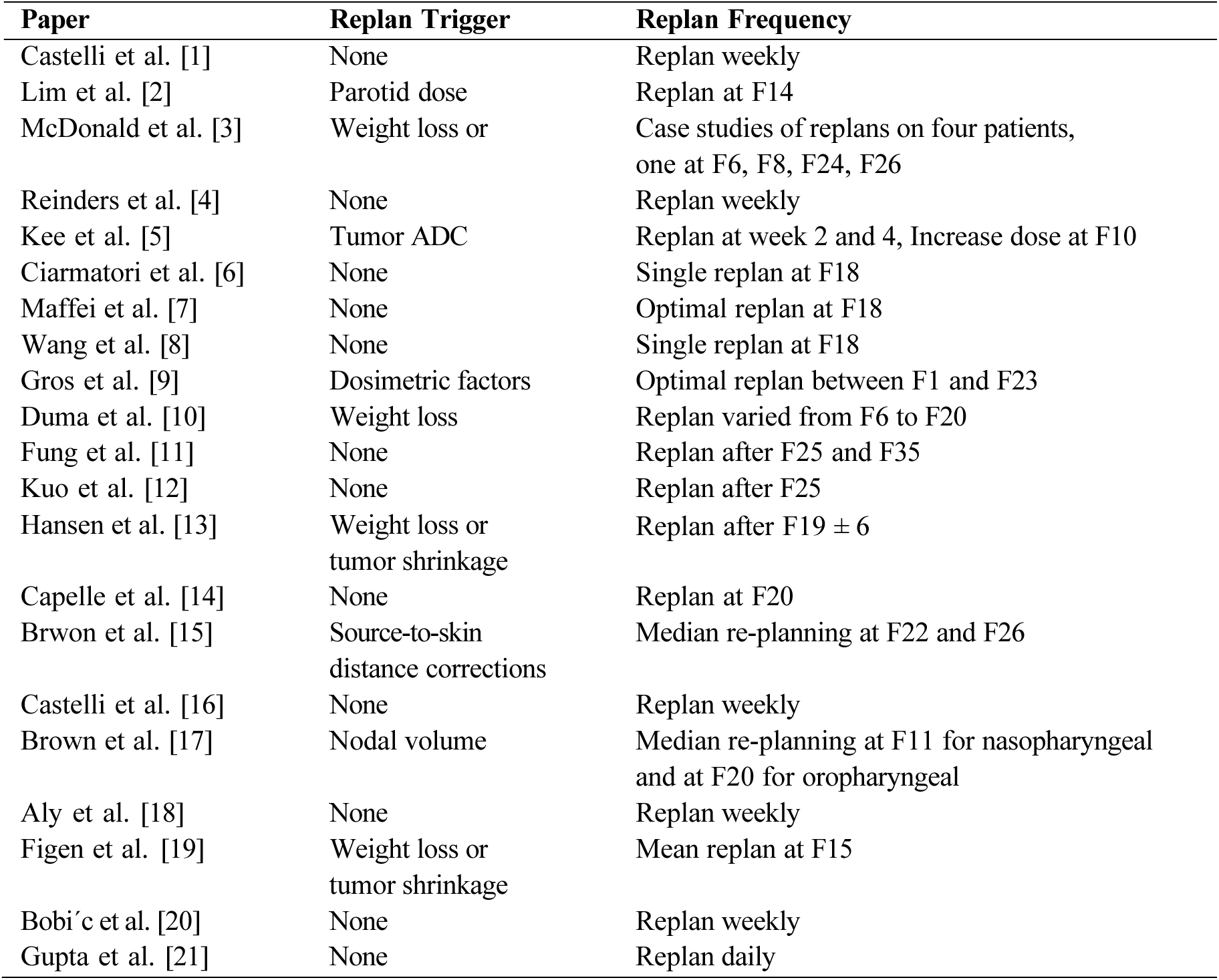
A comprehensive list of relevant papers on head-and-neck-cancer ART along with the number of re-plan and the reported fractions.

| Paper | Replan Trigger | Replan Frequency |
| --- | --- | --- |
| Castelli et al. [1] | None | Replan weekly |
| Lim et al. [2] | Parotid dose | Replan at F14 |
| McDonald et al. [3] | Weight loss or | Case studies of replans on four patients,<br>one at F6, F8, F24, F26 |
| Reinders et al. [4] | None | Replan weekly |
| Kee et al. [5] | Tumor ADC | Replan at week 2 and 4, Increase dose at F10 |
| Ciarmatori et al. [6] | None | Single replan at F18 |
| Maffei et al. [7] | None | Optimal replan at F18 |
| Wang et al. [8] | None | Single replan at F18 |
| Gros et al. [9] | Dosimetric factors | Optimal replan between F1 and F23 |
| Duma et al. [10] | Weight loss | Replan varied from F6 to F20 |
| Fung et al. [11] | None | Replan after F25 and F35 |
| Kuo et al. [12] | None | Replan after F25 |
| Hansen et al. [13] | Weight loss or<br>tumor shrinkage | Replan after $F19 \pm 6$ |
| Capelle et al. [14] | None | Replan at F20 |
| Brwon et al. [15] | Source-to-skin<br>distance corrections | Median re-planning at F22 and F26 |
| Castelli et al. [16] | None | Replan weekly |
| Brown et al. [17] | Nodal volume | Median re-planning at F11 for nasopharyngeal<br>and at F20 for oropharyngeal |
| Aly et al. [18] | None | Replan weekly |
| Figen et al. [19] | Weight loss or<br>tumor shrinkage | Mean replan at F15 |
| Bobić et al. [20] | None | Replan weekly |
| Gupta et al. [21] | None | Replan daily |

## Supplementary Material B. Patient Characteristics

**Table B1:**
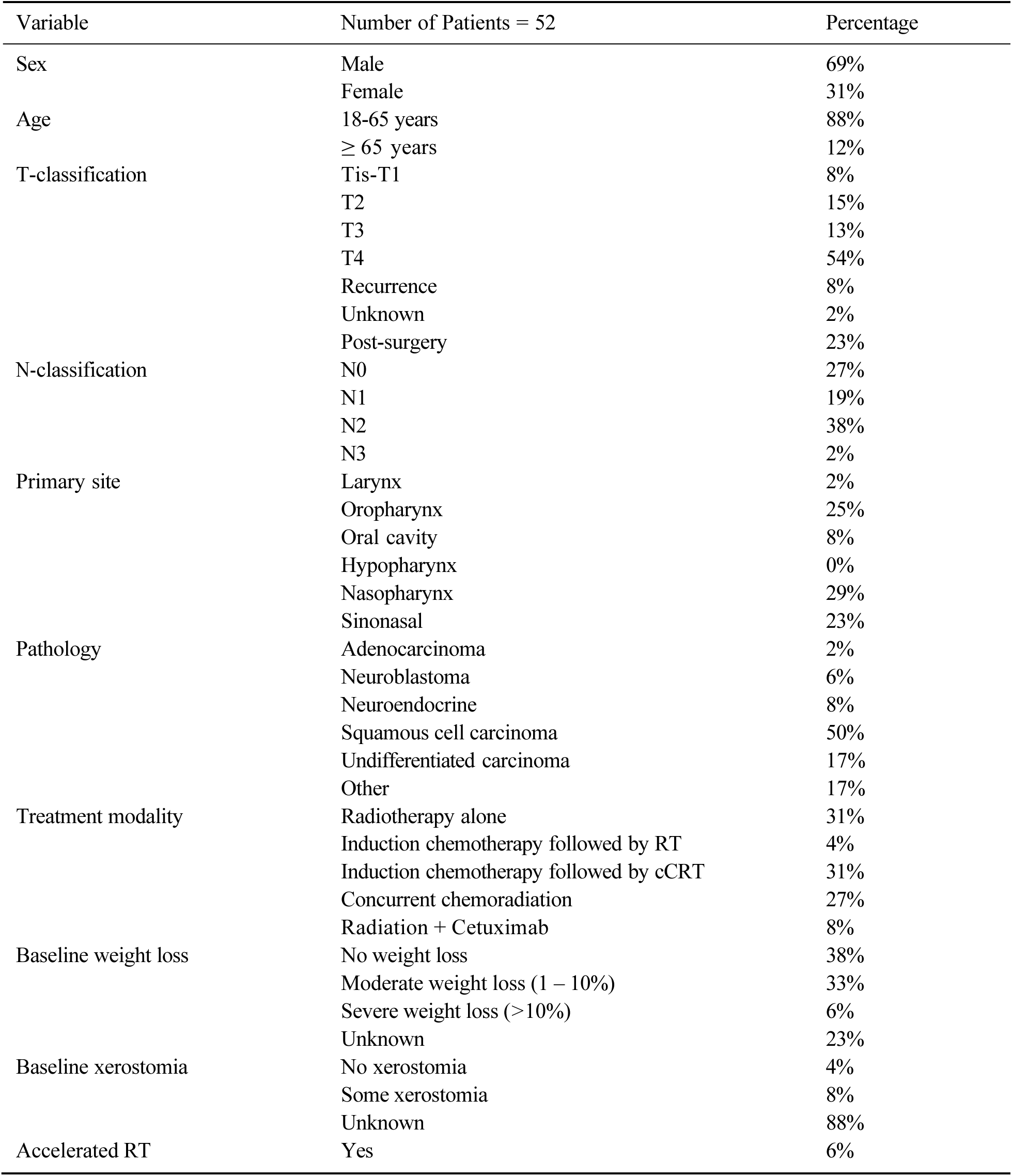
Patient characteristics. Abbreviations: cCRT = concurrent chemoradiation. RT = radiotherapy. TNM classification according to version 3. Accelerated RT: 2 Gy per fraction, 6 times per week.

## Supplementary Material C. Transition Probabilities

### Action 1: No re-planning

The natural transition probabilities from fraction 0 (pre-treatment) to fraction 10 were directly estimated using the information provided in Table 1. For each ΔNTCP category, the ratio of the corresponding number of patients at fraction 10 to the total of 52 patients was taken as the associated transition probability. We note that all patients exhibit ΔNTCP = 0% at fraction 0. These probabilities apply only to the states with the same number of remaining re-plans *b*, by the definition of the ‘no re-planning’ action. The transition probabilities are presented in Supplementary Table C1.

**Table C1:**
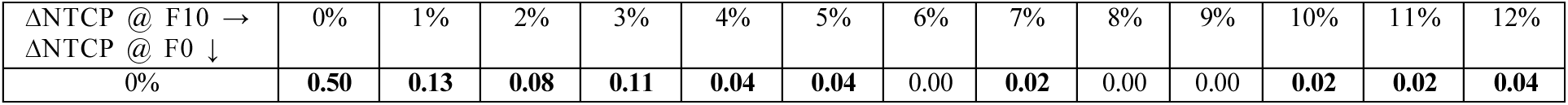
Transition probabilities from fraction 0 (F0) to fraction 10 (F10) under ‘no re-planning.’

The natural transition probabilities from fraction 10 to fraction 15 were estimated in a similar manner. As the number of patients in each ΔNTCP category at fraction 15 were aggregated, a mapping from ΔNTCP categories at fraction 10 to those of fraction 15 was established. To ensure a feasible map, it was assumed that ΔNTCPs may slightly improve (1%) on its own, without intervention. The necessity of this assumption may be observed by the number of patients with ΔNTCP = 12% at Fraction 10 and 15 in Table 1; two patients exhibited ΔNTCP = 12% at fraction 10, while this number at fraction 15 was one patient. As no patient showed ΔNTCP = 6%, 8%, 9% at fraction 10, it was assumed that the patients in these categories retain the same ΔNTCP at fraction 15 with probability 1. The transition probabilities are presented in Supplementary Table C2.

**Table C2:** Transition probabilities from fraction 10 (F10) to fraction 15 (F15) under ‘no re-planning.’ The same probabilities apply to subsequent transitions, i.e., from F15 to F20, from F20 to F25, and from F25 to end-treatment, under ‘no re-planning.’

| $\Delta\text{NTCP @ F15} \rightarrow$<br>$\Delta\text{NTCP @ F10} \downarrow$ | 0% | 1% | 2% | 3% | 4% | 5% | 6% | 7% | 8% | 9% | 10% | 11% | 12% |
| --- | --- | --- | --- | --- | --- | --- | --- | --- | --- | --- | --- | --- | --- |
| 0% | **0.88** | 0.00 | **0.12** | 0.00 | 0.00 | 0.00 | 0.00 | 0.00 | 0.00 | 0.00 | 0.00 | 0.00 | 0.00 |
| 1% | 0.00 | **1.00** | 0.00 | 0.00 | 0.00 | 0.00 | 0.00 | 0.00 | 0.00 | 0.00 | 0.00 | 0.00 | 0.00 |
| 2% | 0.00 | **0.25** | **0.75** | 0.00 | 0.00 | 0.00 | 0.00 | 0.00 | 0.00 | 0.00 | 0.00 | 0.00 | 0.00 |
| 3% | 0.00 | 0.00 | **0.50** | **0.33** | **0.17** | 0.00 | 0.00 | 0.00 | 0.00 | 0.00 | 0.00 | 0.00 | 0.00 |
| 4% | 0.00 | 0.00 | 0.00 | 0.00 | **0.50** | 0.00 | **0.50** | 0.00 | 0.00 | 0.00 | 0.00 | 0.00 | 0.00 |
| 5% | 0.00 | 0.00 | 0.00 | 0.00 | 0.00 | **0.50** | 0.00 | **0.50** | 0.00 | 0.00 | 0.00 | 0.00 | 0.00 |
| 6% | 0.00 | 0.00 | 0.00 | 0.00 | 0.00 | 0.00 | **1.00** | 0.00 | 0.00 | 0.00 | 0.00 | 0.00 | 0.00 |
| 7% | 0.00 | 0.00 | 0.00 | 0.00 | 0.00 | 0.00 | 0.00 | **1.00** | 0.00 | 0.00 | 0.00 | 0.00 | 0.00 |
| 8% | 0.00 | 0.00 | 0.00 | 0.00 | 0.00 | 0.00 | 0.00 | 0.00 | **1.00** | 0.00 | 0.00 | 0.00 | 0.00 |
| 9% | 0.00 | 0.00 | 0.00 | 0.00 | 0.00 | 0.00 | 0.00 | 0.00 | 0.00 | **1.00** | 0.00 | 0.00 | 0.00 |
| 10% | 0.00 | 0.00 | 0.00 | 0.00 | 0.00 | 0.00 | 0.00 | 0.00 | 0.00 | 0.00 | **1.00** | 0.00 | 0.00 |
| 11% | 0.00 | 0.00 | 0.00 | 0.00 | 0.00 | 0.00 | 0.00 | 0.00 | 0.00 | 0.00 | **1.00** | 0.00 | 0.00 |
| 12% | 0.00 | 0.00 | 0.00 | 0.00 | 0.00 | 0.00 | 0.00 | 0.00 | 0.00 | 0.00 | 0.00 | **0.50** | **0.50** |

**Table C3:** Smoothed Transition probabilities from fraction 10 (F10) to fraction 15 (F15) under ‘no re- planning.’ The same probabilities apply to subsequent transitions, i.e., from F15 to F20, from F20 to F25, and from F25 to end- treatment, under ‘no re-planning.’

| $\Delta\text{NTCP @ F15} \rightarrow$<br>$\Delta\text{NTCP @ F10} \downarrow$ | 0% | 1% | 2% | 3% | 4% | 5% | 6% | 7% | 8% | 9% | 10% | 11% | 12% |
| --- | --- | --- | --- | --- | --- | --- | --- | --- | --- | --- | --- | --- | --- |
| 0% | **0.81** | **0.08** | **0.11** | 0.00 | 0.00 | 0.00 | 0.00 | 0.00 | 0.00 | 0.00 | 0.00 | 0.00 | 0.00 |
| 1% | **0.07** | **0.86** | **0.07** | 0.00 | 0.00 | 0.00 | 0.00 | 0.00 | 0.00 | 0.00 | 0.00 | 0.00 | 0.00 |
| 2% | **0.01** | **0.28** | **0.66** | **0.04** | **0.01** | 0.00 | 0.00 | 0.00 | 0.00 | 0.00 | 0.00 | 0.00 | 0.00 |
| 3% | 0.00 | **0.03** | **0.46** | **0.31** | **0.17** | **0.01** | **0.02** | 0.00 | 0.00 | 0.00 | 0.00 | 0.00 | 0.00 |
| 4% | 0.00 | 0.00 | **0.02** | **0.03** | **0.43** | **0.06** | **0.42** | **0.04** | 0.00 | 0.00 | 0.00 | 0.00 | 0.00 |
| 5% | 0.00 | 0.00 | 0.00 | 0.00 | **0.04** | **0.43** | **0.09** | **0.42** | **0.02** | 0.00 | 0.00 | 0.00 | 0.00 |
| 6% | 0.00 | 0.00 | 0.00 | 0.00 | 0.00 | **0.06** | **0.85** | **0.09** | 0.00 | 0.00 | 0.00 | 0.00 | 0.00 |
| 7% | 0.00 | 0.00 | 0.00 | 0.00 | 0.00 | 0.00 | **0.07** | **0.85** | **0.08** | 0.00 | 0.00 | 0.00 | 0.00 |
| 8% | 0.00 | 0.00 | 0.00 | 0.00 | 0.00 | 0.00 | 0.00 | **0.07** | **0.85** | **0.08** | 0.00 | 0.00 | 0.00 |
| 9% | 0.00 | 0.00 | 0.00 | 0.00 | 0.00 | 0.00 | 0.00 | 0.00 | **0.07** | **0.85** | **0.08** | 0.00 | 0.00 |
| 10% | 0.00 | 0.00 | 0.00 | 0.00 | 0.00 | 0.00 | 0.00 | 0.00 | 0.00 | **0.08** | **0.88** | **0.04** | 0.00 |
| 11% | 0.00 | 0.00 | 0.00 | 0.00 | 0.00 | 0.00 | 0.00 | 0.00 | 0.00 | **0.04** | **0.88** | **0.06** | **0.02** |
| 12% | 0.00 | 0.00 | 0.00 | 0.00 | 0.00 | 0.00 | 0.00 | 0.00 | 0.00 | 0.00 | **0.06** | **0.46** | **0.48** |

### Action 2: Re-planning

The impact of ‘re-planning’ on transition probabilities at a specific fraction was modeled by presuming an immediate reduction in ΔNTCP to a proportion relative to the elapsed treatment time, followed by a natural transition governed by the probabilities associated with the reduced ΔNTCP. For instance, ‘re-planning’ at fraction 10 when ΔNTCP is at 5% results in an immediate decrease to ΔNTCP = 2%, reflecting that about one-third of the treatment has been completed. This is then followed by a transition (from F10 to F15) according to the probabilities associated with ΔNTCP = 2% @ F10 in Supplementary Table C2. These probabilities are presented in the following tables. We note that the earliest decision epoch for ‘re-planning’ is fraction 10.

**Table C4:** Transition probabilities from fraction 10 (F10) to fraction 15 (F15) under ‘re-planning.’

| $\Delta\text{NTCP @ F15} \rightarrow$<br>$\Delta\text{NTCP @ F10} \downarrow$ | 0% | 1% | 2% | 3% | 4% | 5% | 6% | 7% | 8% | 9% | 10% | 11% | 12% |
| --- | --- | --- | --- | --- | --- | --- | --- | --- | --- | --- | --- | --- | --- |
| 0% | **0.81** | **0.08** | **0.11** | 0.00 | 0.00 | 0.00 | 0.00 | 0.00 | 0.00 | 0.00 | 0.00 | 0.00 | 0.00 |
| 1% | **0.81** | **0.08** | **0.11** | 0.00 | 0.00 | 0.00 | 0.00 | 0.00 | 0.00 | 0.00 | 0.00 | 0.00 | 0.00 |
| 2% | **0.07** | **0.86** | **0.07** | 0.00 | 0.00 | 0.00 | 0.00 | 0.00 | 0.00 | 0.00 | 0.00 | 0.00 | 0.00 |
| 3% | **0.07** | **0.86** | **0.07** | 0.00 | 0.00 | 0.00 | 0.00 | 0.00 | 0.00 | 0.00 | 0.00 | 0.00 | 0.00 |
| 4% | **0.07** | **0.86** | **0.07** | 0.00 | 0.00 | 0.00 | 0.00 | 0.00 | 0.00 | 0.00 | 0.00 | 0.00 | 0.00 |
| 5% | **0.01** | **0.28** | **0.66** | **0.04** | **0.01** | 0.00 | 0.00 | 0.00 | 0.00 | 0.00 | 0.00 | 0.00 | 0.00 |
| 6% | **0.01** | **0.28** | **0.66** | **0.04** | **0.01** | 0.00 | 0.00 | 0.00 | 0.00 | 0.00 | 0.00 | 0.00 | 0.00 |
| 7% | **0.01** | **0.28** | **0.66** | **0.04** | **0.01** | 0.00 | 0.00 | 0.00 | 0.00 | 0.00 | 0.00 | 0.00 | 0.00 |
| 8% | 0.00 | **0.03** | **0.46** | **0.31** | **0.17** | **0.01** | **0.02** | 0.00 | 0.00 | 0.00 | 0.00 | 0.00 | 0.00 |
| 9% | 0.00 | **0.03** | **0.46** | **0.31** | **0.17** | **0.01** | **0.02** | 0.00 | 0.00 | 0.00 | 0.00 | 0.00 | 0.00 |
| 10% | 0.00 | **0.03** | **0.46** | **0.31** | **0.17** | **0.01** | **0.02** | 0.00 | 0.00 | 0.00 | 0.00 | 0.00 | 0.00 |
| 11% | 0.00 | 0.00 | **0.02** | **0.03** | **0.43** | **0.06** | **0.42** | **0.04** | 0.00 | 0.00 | 0.00 | 0.00 | 0.00 |
| 12% | 0.00 | 0.00 | **0.02** | **0.03** | **0.43** | **0.06** | **0.42** | **0.04** | 0.00 | 0.00 | 0.00 | 0.00 | 0.00 |

**Table C5:** Transition probabilities from fraction 15 (F15) to fraction 20 (F20) under ‘re-planning.’

| $\Delta\text{NTCP @ F20} \rightarrow$<br>$\Delta\text{NTCP @ F15} \downarrow$ | 0% | 1% | 2% | 3% | 4% | 5% | 6% | 7% | 8% | 9% | 10% | 11% | 12% |
| --- | --- | --- | --- | --- | --- | --- | --- | --- | --- | --- | --- | --- | --- |
| 0% | **0.81** | **0.08** | **0.11** | 0.00 | 0.00 | 0.00 | 0.00 | 0.00 | 0.00 | 0.00 | 0.00 | 0.00 | 0.00 |
| 1% | **0.07** | **0.86** | **0.07** | 0.00 | 0.00 | 0.00 | 0.00 | 0.00 | 0.00 | 0.00 | 0.00 | 0.00 | 0.00 |
| 2% | **0.07** | **0.86** | **0.07** | 0.00 | 0.00 | 0.00 | 0.00 | 0.00 | 0.00 | 0.00 | 0.00 | 0.00 | 0.00 |
| 3% | **0.01** | **0.28** | **0.66** | **0.04** | **0.01** | 0.00 | 0.00 | 0.00 | 0.00 | 0.00 | 0.00 | 0.00 | 0.00 |
| 4% | **0.01** | **0.28** | **0.66** | **0.04** | **0.01** | 0.00 | 0.00 | 0.00 | 0.00 | 0.00 | 0.00 | 0.00 | 0.00 |
| 5% | 0.00 | **0.03** | **0.46** | **0.31** | **0.17** | **0.01** | **0.02** | 0.00 | 0.00 | 0.00 | 0.00 | 0.00 | 0.00 |
| 6% | 0.00 | **0.03** | **0.46** | **0.31** | **0.17** | **0.01** | **0.02** | 0.00 | 0.00 | 0.00 | 0.00 | 0.00 | 0.00 |
| 7% | 0.00 | 0.00 | **0.02** | **0.03** | **0.43** | **0.06** | **0.42** | **0.04** | 0.00 | 0.00 | 0.00 | 0.00 | 0.00 |
| 8% | 0.00 | 0.00 | **0.02** | **0.03** | **0.43** | **0.06** | **0.42** | **0.04** | 0.00 | 0.00 | 0.00 | 0.00 | 0.00 |
| 9% | 0.00 | 0.00 | 0.00 | 0.00 | **0.04** | **0.43** | **0.09** | **0.42** | **0.02** | 0.00 | 0.00 | 0.00 | 0.00 |
| 10% | 0.00 | 0.00 | 0.00 | 0.00 | **0.04** | **0.43** | **0.09** | **0.42** | **0.02** | 0.00 | 0.00 | 0.00 | 0.00 |
| 11% | 0.00 | 0.00 | 0.00 | 0.00 | 0.00 | **0.06** | **0.85** | **0.09** | 0.00 | 0.00 | 0.00 | 0.00 | 0.00 |
| 12% | 0.00 | 0.00 | 0.00 | 0.00 | 0.00 | **0.06** | **0.85** | **0.09** | 0.00 | 0.00 | 0.00 | 0.00 | 0.00 |

**Table C6:**
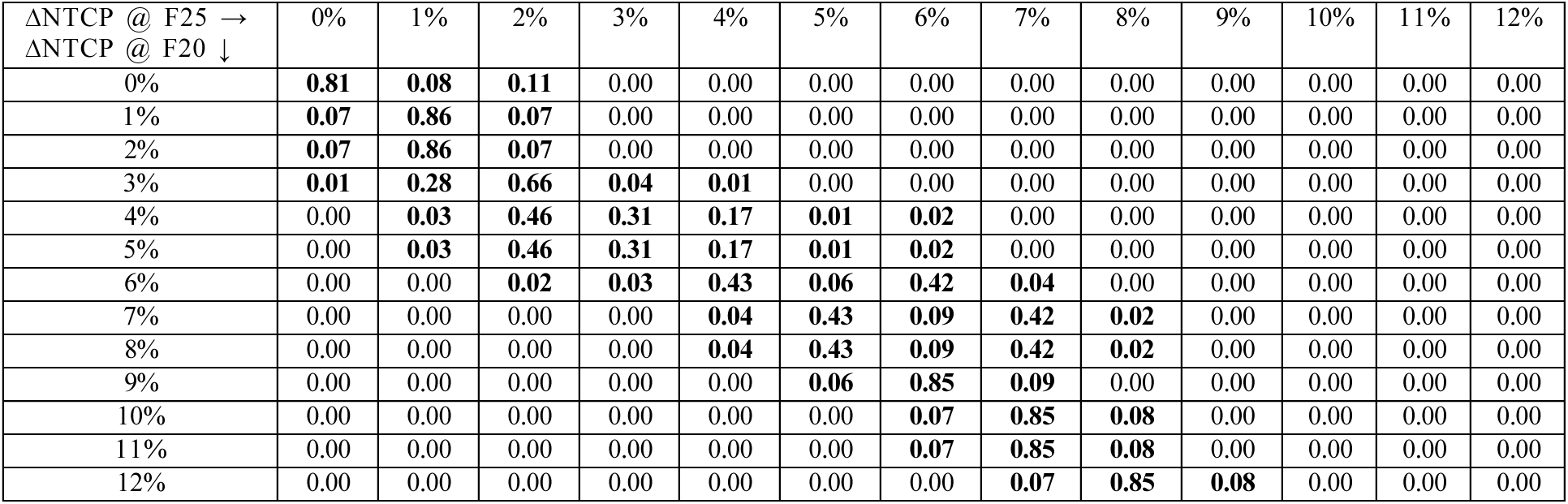
Transition probabilities from fraction 20 (F20) to fraction 25 (F25) under ‘re-planning.’

| $\Delta\text{NTCP @ F25} \rightarrow$<br>$\Delta\text{NTCP @ F20} \downarrow$ | 0% | 1% | 2% | 3% | 4% | 5% | 6% | 7% | 8% | 9% | 10% | 11% | 12% |
| --- | --- | --- | --- | --- | --- | --- | --- | --- | --- | --- | --- | --- | --- |
| 0% | **0.81** | **0.08** | **0.11** | 0.00 | 0.00 | 0.00 | 0.00 | 0.00 | 0.00 | 0.00 | 0.00 | 0.00 | 0.00 |
| 1% | **0.07** | **0.86** | **0.07** | 0.00 | 0.00 | 0.00 | 0.00 | 0.00 | 0.00 | 0.00 | 0.00 | 0.00 | 0.00 |
| 2% | **0.07** | **0.86** | **0.07** | 0.00 | 0.00 | 0.00 | 0.00 | 0.00 | 0.00 | 0.00 | 0.00 | 0.00 | 0.00 |
| 3% | **0.01** | **0.28** | **0.66** | **0.04** | **0.01** | 0.00 | 0.00 | 0.00 | 0.00 | 0.00 | 0.00 | 0.00 | 0.00 |
| 4% | 0.00 | **0.03** | **0.46** | **0.31** | **0.17** | **0.01** | **0.02** | 0.00 | 0.00 | 0.00 | 0.00 | 0.00 | 0.00 |
| 5% | 0.00 | **0.03** | **0.46** | **0.31** | **0.17** | **0.01** | **0.02** | 0.00 | 0.00 | 0.00 | 0.00 | 0.00 | 0.00 |
| 6% | 0.00 | 0.00 | **0.02** | **0.03** | **0.43** | **0.06** | **0.42** | **0.04** | 0.00 | 0.00 | 0.00 | 0.00 | 0.00 |
| 7% | 0.00 | 0.00 | 0.00 | 0.00 | **0.04** | **0.43** | **0.09** | **0.42** | **0.02** | 0.00 | 0.00 | 0.00 | 0.00 |
| 8% | 0.00 | 0.00 | 0.00 | 0.00 | **0.04** | **0.43** | **0.09** | **0.42** | **0.02** | 0.00 | 0.00 | 0.00 | 0.00 |
| 9% | 0.00 | 0.00 | 0.00 | 0.00 | 0.00 | **0.06** | **0.85** | **0.09** | 0.00 | 0.00 | 0.00 | 0.00 | 0.00 |
| 10% | 0.00 | 0.00 | 0.00 | 0.00 | 0.00 | 0.00 | **0.07** | **0.85** | **0.08** | 0.00 | 0.00 | 0.00 | 0.00 |
| 11% | 0.00 | 0.00 | 0.00 | 0.00 | 0.00 | 0.00 | **0.07** | **0.85** | **0.08** | 0.00 | 0.00 | 0.00 | 0.00 |
| 12% | 0.00 | 0.00 | 0.00 | 0.00 | 0.00 | 0.00 | 0.00 | **0.07** | **0.85** | **0.08** | 0.00 | 0.00 | 0.00 |

**Table C7:**
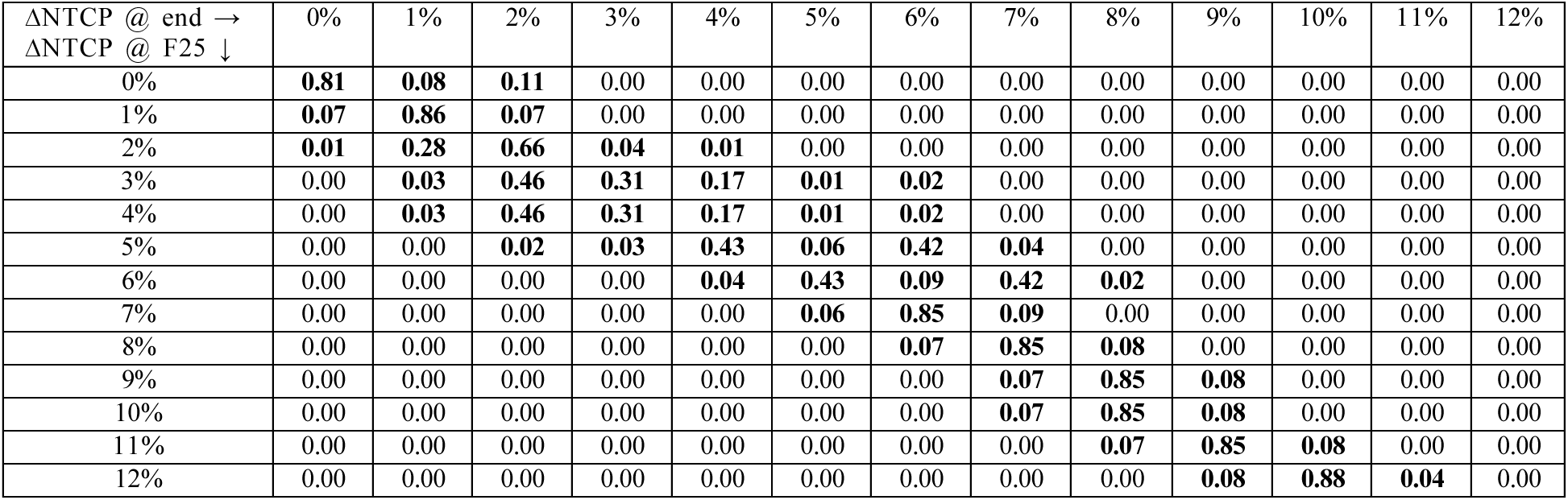
Transition probabilities from fraction 25 (F25) to end-treatment under ‘re-planning.’

| $\Delta\text{NTCP @ end} \rightarrow$<br>$\Delta\text{NTCP @ F25} \downarrow$ | 0% | 1% | 2% | 3% | 4% | 5% | 6% | 7% | 8% | 9% | 10% | 11% | 12% |
| --- | --- | --- | --- | --- | --- | --- | --- | --- | --- | --- | --- | --- | --- |
| 0% | **0.81** | **0.08** | **0.11** | 0.00 | 0.00 | 0.00 | 0.00 | 0.00 | 0.00 | 0.00 | 0.00 | 0.00 | 0.00 |
| 1% | **0.07** | **0.86** | **0.07** | 0.00 | 0.00 | 0.00 | 0.00 | 0.00 | 0.00 | 0.00 | 0.00 | 0.00 | 0.00 |
| 2% | **0.01** | **0.28** | **0.66** | **0.04** | **0.01** | 0.00 | 0.00 | 0.00 | 0.00 | 0.00 | 0.00 | 0.00 | 0.00 |
| 3% | 0.00 | **0.03** | **0.46** | **0.31** | **0.17** | **0.01** | **0.02** | 0.00 | 0.00 | 0.00 | 0.00 | 0.00 | 0.00 |
| 4% | 0.00 | **0.03** | **0.46** | **0.31** | **0.17** | **0.01** | **0.02** | 0.00 | 0.00 | 0.00 | 0.00 | 0.00 | 0.00 |
| 5% | 0.00 | 0.00 | **0.02** | **0.03** | **0.43** | **0.06** | **0.42** | **0.04** | 0.00 | 0.00 | 0.00 | 0.00 | 0.00 |
| 6% | 0.00 | 0.00 | 0.00 | 0.00 | **0.04** | **0.43** | **0.09** | **0.42** | **0.02** | 0.00 | 0.00 | 0.00 | 0.00 |
| 7% | 0.00 | 0.00 | 0.00 | 0.00 | 0.00 | **0.06** | **0.85** | **0.09** | 0.00 | 0.00 | 0.00 | 0.00 | 0.00 |
| 8% | 0.00 | 0.00 | 0.00 | 0.00 | 0.00 | 0.00 | **0.07** | **0.85** | **0.08** | 0.00 | 0.00 | 0.00 | 0.00 |
| 9% | 0.00 | 0.00 | 0.00 | 0.00 | 0.00 | 0.00 | 0.00 | **0.07** | **0.85** | **0.08** | 0.00 | 0.00 | 0.00 |
| 10% | 0.00 | 0.00 | 0.00 | 0.00 | 0.00 | 0.00 | 0.00 | **0.07** | **0.85** | **0.08** | 0.00 | 0.00 | 0.00 |
| 11% | 0.00 | 0.00 | 0.00 | 0.00 | 0.00 | 0.00 | 0.00 | 0.00 | **0.07** | **0.85** | **0.08** | 0.00 | 0.00 |
| 12% | 0.00 | 0.00 | 0.00 | 0.00 | 0.00 | 0.00 | 0.00 | 0.00 | 0.00 | **0.08** | **0.88** | **0.04** | 0.00 |

## Supplementary Material D. Optimal ΔNTCP Thresholds for Re-planning

**Fig. D1:**
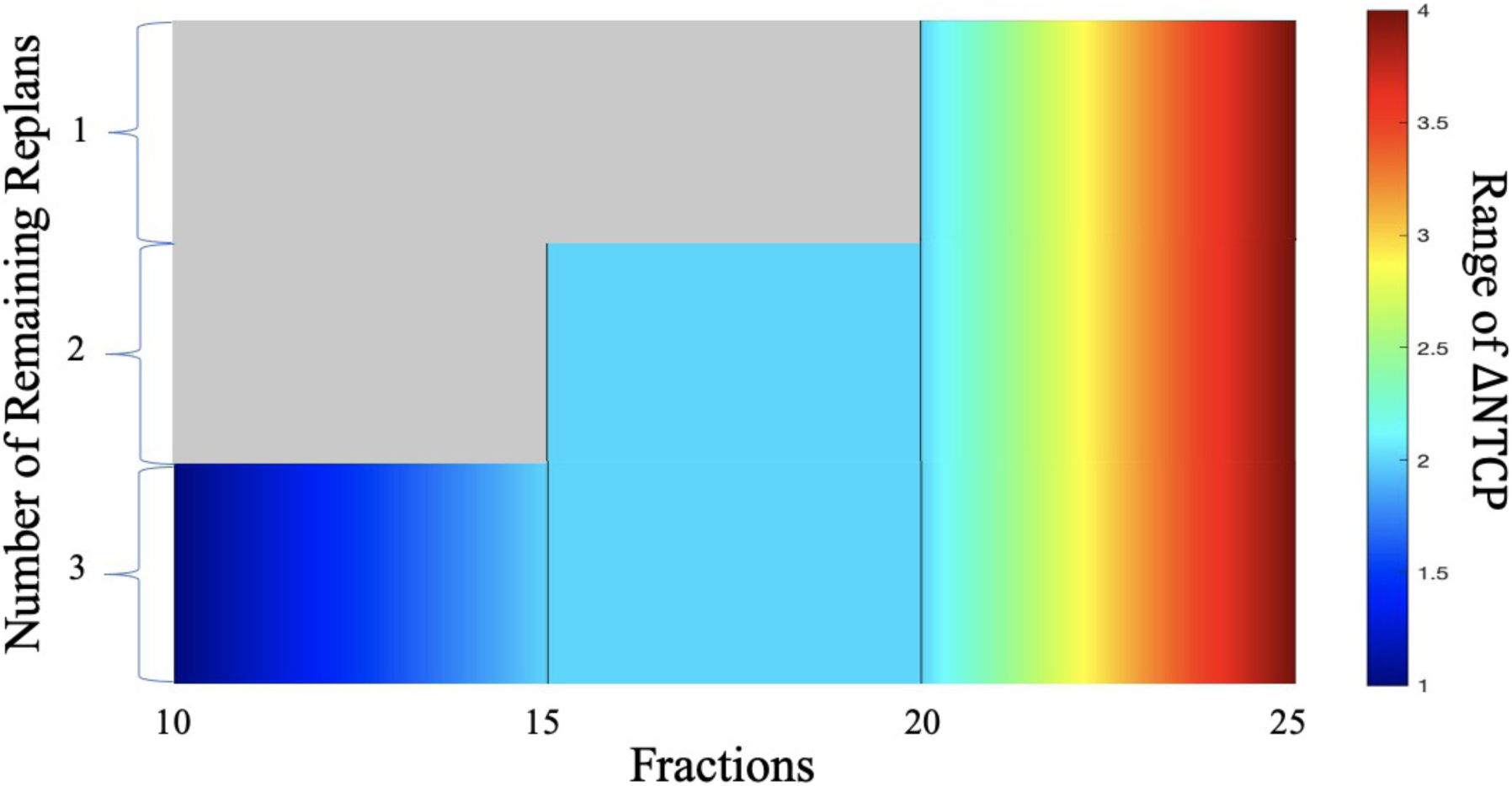
Optimal ΔNTCP thresholds for re-planning for *B* = 3. Optimal ΔNTCP thresholds for re-planning at fractions 10, 15, 20, and 25 are 1%, 2%, 2%, and 4%, respectively. The optimal ΔNTCP thresholds increase with respect to fraction but remain constant with respect to the number of replans remaining.

## Supplementary Material E. NTCP Models

**Table E1:**
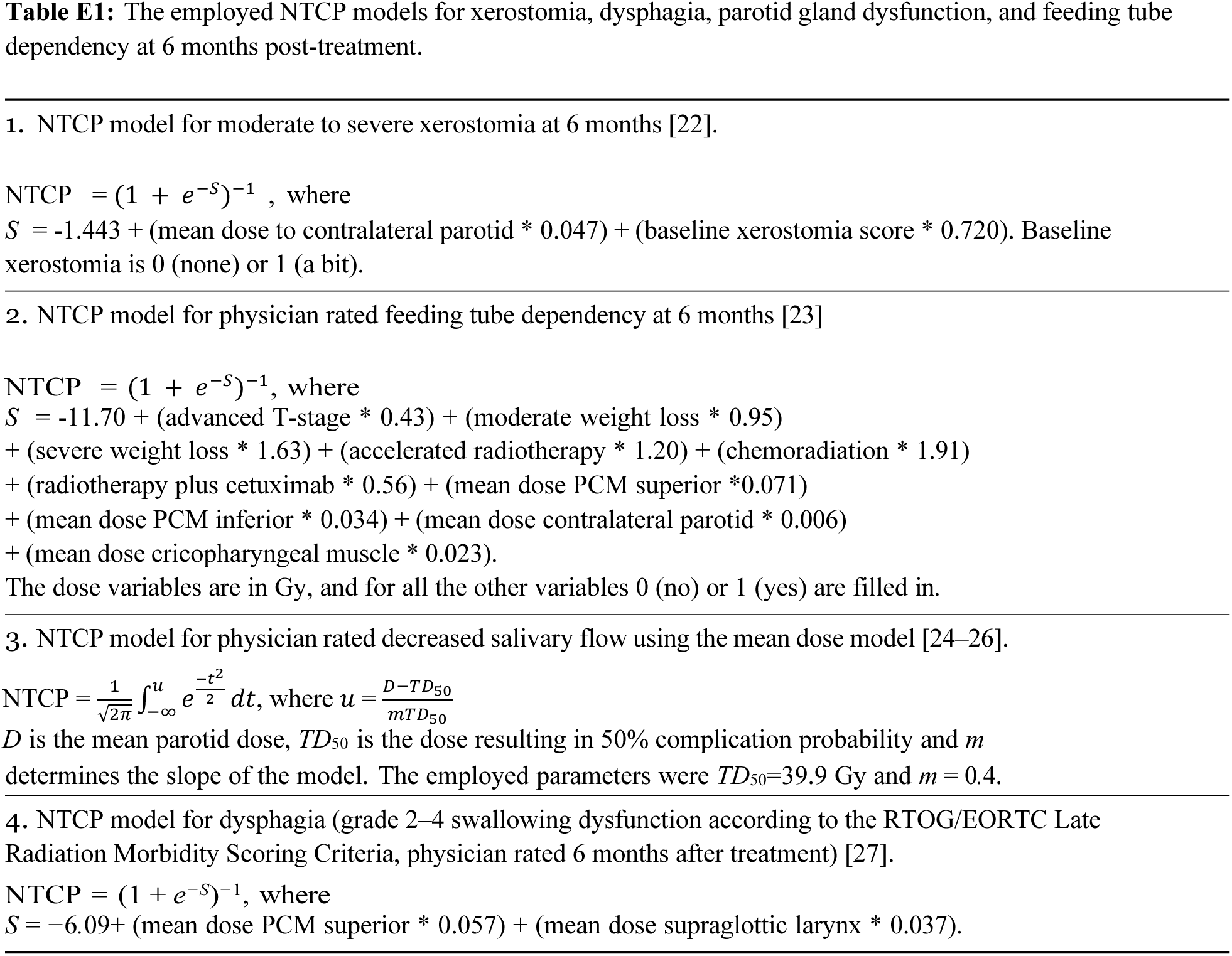
The employed NTCP models for xerostomia, dysphagia, parotid gland dysfunction, and feeding tube dependency at 6 months post-treatment.

## References

1. Iacovelli NA, et al. Role of IMRT/VMAT-based dose and volume parameters in predicting 5-year local control and survival in nasopharyngeal cancer patients. Frontiers in Oncology 2020;10:e518110.

2. Barker Jr JL, et al. Quantification of volumetric and geometric changes occurring during fractionated radiotherapy for head-and-neck cancer using an integrated CT/linear accelerator system. International Journal of Radiation Oncology, Biology, Physics 2004;59(4):960–70.

3. Beltran M, Ramos M, Rovira JJ, Perez-Hoyos S, Sancho M, Puertas E, Benavente S, Ginjaume M, Giralt J. Dose variations in tumor volumes and organs at risk during IMRT for head-and-neck cancer. Journal of Applied Clinical Medical Physics 2012;13(6):e3723.

4. Chen W, Bai P, Pan J, Xu Y, Chen K. Changes in tumor volumes and spatial locations relative to normal tissues during cervical cancer radiotherapy assessed by cone beam computed tomography. Technology in Cancer Research and Treatment 2017;16(2):246–52.

5. Beadle BM, Chan AW. The potential of adaptive radiotherapy for patients with head and neck cancer-too much or not enough? JAMA Oncology 2023; 9(8):1064–65.

6. Castelli J, Simon A, Lafond C, Perichon N, Rigaud B, Chajon E, De Bari B, Ozsahin M, Bourhis J, de Crevoisier R. Adaptive radiotherapy for head and neck cancer. Acta Oncology 2018;57(10):1284–92.

7. Figen M, Öksüz DÇ, Duman E, Prestwich R, Dyker K, Cardale K, Ramasamy S, Murray P, Şen M. Radiotherapy for head and neck cancer: Evaluation of triggered adaptive replanning in routine practice. Frontiers in Oncology 2020;59(4):960–70.

8. Håkansson K, Giannoulis E, Lindegaard A, Friborg J, Vogelius I. CBCT-based online adaptive radiotherapy for head and neck cancer-dosimetric evaluation of first clinical. Acta Oncologica 2023; 62(11):1369–1374.

9. Lavrova E, Garrett MD, Wang YF, Chin C, Elliston C, Savacool M, Price M, Kachnic LA, Horowitz DP. Adaptive radiation therapy: A review of CT-based techniques. Radiology. Imaging Cancer 2023; 5(4):e230011.

10. O’Hara CJ, Bird D, Al-Qaisieh B, Speight R. Assessment of CBCT-based synthetic CT generation accuracy for adaptive radiotherapy planning. Journal of Applied Clinical Medical Physics 2022; 23(11):e13737.

11. Brock KK. Adaptive radiotherapy: Moving into the future. Seminars in Radiation Oncology 2019; 29(3):181–184.

12. Wahid, KA et al. Harnessing uncertainty in radiotherapy auto-segmentation quality assurance. Physics and Imaging in Radiation Oncology 2023; 29:100526.

13. Allen C, Yeo AU, Hardcastle N, Franich RD. Evaluating synthetic computed tomography images for adaptive radiotherapy decision making in head and neck cancer. Physics and Imaging in Radiation Oncology 2023; 27:100478.

14. Taasti VT, et al. Clinical evaluation of synthetic computed tomography methods in adaptive proton therapy of lung cancer patients. Physics and Imaging in Radiation Oncology 2023;27:e100459.

15. Huynh E, Hosny A, Guthier C, Bitterman SF, Hass-Kogan DA, Kann B, Aerts HJWL, Mak RG. Artificial intelligence in radiation oncology. Nature Reviews Clinical Oncology 2020; 17:771–81.

16. van de Schoot AJ, Hoffmans D, van Ingen KM, Simons MJ, Wiersma J. Characterization of Ethos therapy systems for adaptive radiation therapy: A multi- machine comparison. Journal of Applied Clinical Medical Physics 2023;24:e13905.

17. Varian Ethos. https://www.varian.com; 2024 [accessed 24 February 2024].

18. Avgousti R, Antypas C, Armpilia C, Simopoulou F, Liakouli Z, Karaiskos P, Kouloulias V, Kyrodimos E, Moulopoulos LA, Zygogianni A. Adaptive radiation therapy: When, how and what are the benefits that literature provides? Cancer Radiotherapie: Journal de la Societe Francaise de Radiotherapie Oncologique 2022;26(4):622–36.

19. Bahig H, et al. Magnetic resonance-based response assessment and dose adaptation in human papilloma virus positive tumors of the oropharynx treated with radiotherapy (MR-ADAPTOR): An R-IDEAL stage 2a-2b/Bayesian phase II trial. Clinical and Translational Radiation Oncology 2018;13:19–23.

20. Castelli J, et al. Weekly adaptive radiotherapy vs standard intensity-modulated radiotherapy for improving salivary function in patients with head and neck cancer: A phase 3 randomized clinical trial. JAMA Oncology 2023;9(8):1056–64.

21. Westerhoff JM, et al. Safety and tolerability of online adaptive high-field magnetic resonance– guided radiotherapy. JAMA Network Open 2024;7(5):e2410819.

22. Heukelom J, et al. Differences between planned and delivered dose for head and neck cancer, and their consequences for normal tissue complication probability and treatment adaptation. Radiotherapy and Oncology 2020;142:100–6.

23. Feinberg EA, Shwartz A. Handbook of Markov Decision Processes: Methods and Applications. New York: Springer; 2002.

24. Puterman ML. Markov Decision Processes: Discrete Stochastic Dynamic Programming. New York: Wiley-Interscience; 2005.

25. Beck JR, Pauker SG. The Markov process in medical prognosis. Medical Decision Making 1983;3(4):419–58.

26. Kuntz KM, Russell LB, Owens DK, Sanders GD, Trikalinos TA, Salomon JA. Decision models in cost-effectiveness analysis. Cost-Effectiveness in Health and Medicine. New York: Oxford University Press, 2016.

27. Ng SP et al. Surveillance imaging for patients with head and neck cancer treated with definitive radiotherapy: A partially observed Markov decision process model. Cancer 2020;126(4):749–56.

28. Alagoz O, Hsu H, Schaefer AJ, Roberts MS. Markov decision processes: A tool for sequential decision making under uncertainty. Medical Decision Making: An International Journal of the Society for Medical Decision Making 2010;30(4):474–83.

29. Alagoz O, Maillart LM, Schaefer AJ, Roberts MS. The optimal timing of living-donor liver transplantation. Management Science 2004;50(10):1420–30.

30. Denton BT, Kurt M, Shah ND, Bryant SC, Smith SA. Optimizing the start time of statin therapy for patients with diabetes. Medical Decision Making 2009;29(3):351–67.

31. Shechter SM, Bailey MD, Schaefer AJ, Roberts MS. The optimal time to initiate HIV therapy under ordered health states. Operations Research 2008;56(1):20–33.

32. MATLAB version: 9.13.0 (R2022b), Natick, Massachusetts: The MathWorks Inc.; 2022.

33. Zhang L, Dong L, Court L, Wang H, Gillin M, Mohan R. TU-EE-A4-05: Validation of CT- assisted targeting (CAT) software for soft tissue and bony target localization. Medical Physics 2005;32:2106.

34. Maki RG, et al. Phase II study of sorafenib in patients with metastatic or recurrent sarcomas. Journal of Clinical Oncology: Official Journal of the American Society of Clinical Oncology 2009;27(19):3133–40.

35. Stieb S, Lee A, van Dijk LV, Frank S, Fuller CD, Blanchard P. NTCP modeling of late effects for head and neck cancer: A systematic review. International Journal of Particle Therapy 2021;8(1):95–107.

36. Hosseinian S, Hemmati M, Dede C, Salzillo TC, van Dijk LV, Mohamed ASR, Lai SY, Schaefer AJ, Fuller CD. Cluster-based toxicity estimation of osteoradionecrosis via unsupervised machine learning: Moving beyond single dose-parameter normal tissue complication probability by using whole dose-volume histograms for cohort risk stratification. International Journal of Radiation Oncology, Biology, Physics 2024;119(5):1569–78.

37. American Cancer Society. Head and Neck Cancer: Statistics. https://www.cancer.net/cancer-types/head-and-neck-cancer/statistics; 2024 [accessed 24 February 2024].

38. Gan Y, Langendijk JA, van der Schaaf A, van den Bosch L, Oldehinkel E, Lin Z, Both S, Brouwer CL. An efficient strategy to select head and neck cancer patients for adaptive radiotherapy. Radiotherapy and Oncology: Journal of the European Society for Therapeutic Radiology and Oncology 2023;186:e109763.

39. Gan Y, Langendijk JA, Oldehinkel E, Lin Z, Both S, Brouwer CL. Optimal timing of re-planning for head and neck adaptive radiotherapy. Journal of the European Society for Therapeutic Radiology and Oncology 2024;194:e110145.

40. Heukelom J, et al. Adaptive and innovative radiation treatment for improving cancer treatment outcome (ARTFORCE); a randomized controlled phase II trial for individualized treatment of head and neck cancer. BMC Cancer 2013;13:84.

41. Leeuw AD, et al. Acute toxicity in ARTFORCE: A randomized phase III dose-painting trial in head and neck cancer. International Journal of Radiation Oncology, Biology, Physics 2022;114(3):S98.

42. Mohamad I, et al. The evolving role of stereotactic body radiation therapy for head and neck cancer: Where do we stand? Cancers (Basel) 2023;15(20):e5010.

43. Lalonde A, Bobić M, Sharp GC, Chamseddine I, Winey B, Paganetti H. Evaluating the effect of setup uncertainty reduction and adaptation to geometric changes on normal tissue complication probability using online adaptive head and neck intensity modulated proton therapy. Physics in Medicine and Biology 2023;68(11):e115018.

44. Yap LM, Jamalludin Z, Ng AH, Ung NM. A multi-center survey on adaptive radiation therapy for head and neck cancer in Malaysia. Physical and Engineering Sciences in Medicine 2023;46(3):1331–40.

45. Bertholet J, et al. Patterns of practice for adaptive and real-time radiation therapy (POP-ART RT) part II: Offline and online plan adaption for interfractional changes. Radiotherapy and Oncology: Journal of the European Society for Therapeutic Radiology and Oncology 2020;153:88–96.

## References

[1] Fuller CD. Introduction to Radiation Oncology, 10.6084/m9.figshare.22582207.v1; 2023 [accessed 24 February 2024].

## References

[1] Castelli J, et al. Weekly adaptive radiotherapy vs standard intensity-modulated radiotherapy for improving salivary function in patients with head and neck cancer: A phase 3 randomized clinical trial. JAMA Oncology 2023;9(8):1056–64.

[2] Lim SY, Tran A, Tran ANK, Sobremonte A, Fuller CD, Simmons L, Yang J. Dose accumulation of daily adaptive plans to decide optimal plan adaptation strategy for head-and-neck patients treated with MR-Linac. Medical Dosimetry: Official Journal of the American Association of Medical Dosimetrists 2022;47(1):103–9.

[3] McDonald BA, et al. Initial feasibility and clinical implementation of daily MR- guided adaptive head and neck cancer radiation therapy on a 1.5T MR-Linac system: Prospective R-IDEAL 2a/2b systematic clinical evaluation of technical innovation. International Journal of Radiation Oncology, Biology, Physics 2021;109(5):1606–18.

[4] Reinders FCJ, de Ridder M, Doornaert PAH, Raaijmakers CPJ, Philippens MEP. Individual elective lymph node irradiation for the reduction of complications in head and neck cancer patients (iNode): A phase-I feasibility trial protocol. Clinical and Translational Radiation Oncology 2022;39:e100574.

[5] H. Kee, et al. Optimising radiation therapy in head and neck cancers using functional image-guided radiotherapy and novel biomarkers. https://clinicaltrials.gov/study/NCT04242459; 2021 [accessed 16 September 2024].

[6] Ciarmatori A, Maffei N, Mistretta GM, Ceroni P, Bernabei A, Meduri B, D’Angelo E, Bruni A, Giacobazzi P, Lohr F, Guidi G. Evaluation of the effectiveness of novel single-intervention adaptive radiotherapy strategies based on daily dose accumulation. Medical Dosimetry: Official Journal of the American Association of Medical Dosimetrists 2019;44(4):379–84.

[7] Maffei N, et al. SIS epidemiological model for adaptive RT: Forecasting the parotid glands shrinkage during tomotherapy treatment. Medical physics 2016;43(7):e4294.

[8] X. Wang, J. Lu, X. Xiong, G. Zhu, H. Ying, S. He, W. Hu, and C. Hu. Anatomic and dosimetric changes during the treatment course of intensity-modulated radiotherapy for locally advanced nasopharyngeal carcinoma. Medical Dosimetry: Official Journal of the American Association of Medical Dosimetrists 2010;35(2):151–7.

[9] Gros SAA, Santhanam AP, Block AM, Emami B, Lee BH, Joyce C. Retrospective clinical evaluation of a decision-support software for adaptive radiotherapy of head and neck cancer patients. Frontiers in Oncology 2022;12:e777793.

[10] Duma MN, Kampfer S, Schuster T, Winkler C, Geinitz H. Adaptive radiotherapy for soft tissue changes during helical tomotherapy for head and neck cancer. Strahlentherapie und Onkologie 2012;188(3):243–7.

[11] Fung WW, Wu VW, Teo PM. Dosimetric evaluation of a three-phase adaptive radiotherapy for nasopharyngeal carcinoma using helical tomotherapy. Medical Dosimetry: Official Journal of the American Association of Medical Dosimetrists 2012;37(1):92–7.

[12] Kuo YC, Wu TH, Chung TS, Huang KW, Chao KS, Su WC, Chiou JF. Effect of regression of enlarged neck lymph nodes on radiation doses received by parotid glands during intensity-modulated radiotherapy for head and neck cancer. American Journal of Clinical Oncology 2006;29(6):600–5.

[13] Hansen EK, Bucci MK, Quivey JM, Weinberg V, Xia P. Repeat CT imaging and replanning during the course of IMRT for head-and-neck cancer. International Journal of Radiation Oncology, Biology, Physics 2006;64(2):355–62.

[14] Capelle L, Mackenzie M, Field C, Parliament M, Ghosh S, Scrimger R. Adaptive radiotherapy using helical tomotherapy for head and neck cancer in definitive and postoperative settings: Initial results. Clinical Oncology (Royal College of Radiologists (Great Britain)) 2012;24(3):208–15.

[15] Brown E, Porceddu S, Owen R, Harden F. Developing an adaptive radiotherapy technique for virally mediated head and neck cancer. Journal of Medical Imaging and Radiation Sciences 2013;44(3):134–40.

[16] Castelli J, et al. Impact of head and neck cancer adaptive radiotherapy to spare the parotid glands and decrease the risk of xerostomia. Radiation Oncology 2015;10(6).

[17] Brown E, Owen R, Harden F, Mengersen K, Oestreich K, Houghton W, Poulsen M, Harris S, Lin C, Porceddu S. Head and neck adaptive radiotherapy: Predicting the time to replan. Asia-Pacific Journal of Clinical Oncology 2016;12(4):460–7.

[18] Aly F, Miller AA, Jameson MG, Metcalfe PE. A prospective study of weekly intensity modulated radiation therapy plan adaptation for head and neck cancer: Improved target coverage and organ at risk sparing. Australasian Physical and Engineering Sciences in Medicine 2019;42(1):43–51.

[19] Figen M, Öksüz DÇ, Duman E, Prestwich R, Dyker K, Cardale K, Ramasamy S, Murray P, Şen M. Radiotherapy for head and neck cancer: Evaluation of triggered adaptive replanning in routine practice. Frontiers in Oncology 2020;59(4):960–70.

[20] Bobić M, Lalonde A, Sharp GC, Grassberger C, Verburg JM, Winey BA, Lomax AJ, Paganetti H. Comparison of weekly and daily online adaptation for head and neck intensity-modulated proton therapy. Physics in Medicine and Biology 2021;66(5):10.1088/1361–6560/abe050.

[21] Gupta A, Dunlop A, Mitchell A, McQuaid D, Nill S, Barnes H, Newbold K, Nutting C, Bhide S, Oelfke U, Harrington KJ, Wong KH. Online adaptive radiotherapy for head and neck cancers on the MR linear accelerator: Introducing a novel modified adapt-to-shape approach. Clinical and Translational Radiation Oncology 2021; 32:48–51.

[22] Beetz I. NTCP models for patient-rated xerostomia and sticky saliva after treatment with intensity modulated radiotherapy for head and neck cancer: The role of dosimetric and clinical factors. Radiotherapy and Oncology 2012;105(1):101–6.

[23] Wopken K. Development of a multivariable normal tissue complication probability (NTCP) model for tube feeding dependence after curative radiotherapy/chemo- radiotherapy in head and neck cancer. Radiotherapy and Oncology 2014;113(1):95–101.

[24] Dijkema T, Raaijmakers CP, Ten Haken RK, Roesink JM, Braam PM, Houweling AC, Moerland MA, Eisbruch A, Terhaard CH. Parotid gland function after radiotherapy: The combined Michigan and Utrecht experience. International Journal of Radiation Oncology, Biology, Physics 2010;78(2):449–53.

[25] Lyman JT. Complication probability as assessed from dose-volume histograms. Radiation Research. Supplement 1985;8:S13–9.

[26] Kutcher GJ, Burman C. Calculation of complication probability factors for non- uniform normal tissue irradiation: The effective volume method. International Journal of Radiation Oncology, Biology, Physics 1989;16(6):1623–30.

[27] Christianen MEMC. Predictive modelling for swallowing dysfunction after primary (chemo) radiation: Results of a prospective observational study. Radiotherapy and Oncology: Journal of the European Society for Therapeutic Radiology and Oncology 2012;105(1):107–14.

